# Cell-Free DNA Concentration as a Mutation-Agnostic Readout of Systemic Tumor Burden and Prognosis: A Prospective Analysis of 1,000 Patients

**DOI:** 10.64898/2026.07.27.26359064

**Authors:** Shravan Leonard-Murali, Mathangi Chandramouli, Christopher Sherry, Sefali Patel, Nujsaubnusi Vue, Patricia Petrosko, Phillip H. Gallo, Philip E. Schumacher, Alexander H. Shannon, Casey J. Allen, John M. Nakayama, Thomas W. Rachman, Oana Carja, Russell S. Schwartz, Ali H. Zaidi, William A. LaFramboise, David L. Bartlett, Patrick L. Wagner

## Abstract

**Background:** Objective assessment of tumor burden in patients with solid tumors remains inexact. Furthermore, while mutation-based liquid biopsies are specific, they are limited by tumor heterogeneity and the requirement for detectable clonal mutations. Cell-free DNA (cfDNA) concentration, a cost-effective substrate for mutation-focused liquid biopsy, has shown promise as a “molecular tumor burden” biomarker.

**Methods:** We analyzed cfDNA concentration from 1000 unique cancer patients using standardized protocols. Demographics, AJCC stage, clinical anatomic tumor burden and oncologic outcome variables were collected. Associations were assessed with non-parametric tests. Multivariable linear regression and Cox proportional hazards models were used to identify independent predictors and survival associations.

**Results:** CfDNA concentration varied broadly (median 6.25 ng/mL; range 0.5-1132.9). Anatomic tumor burden, including tumor number (rho=0.25, p<0.0001) and largest tumor diameter (rho=0.25, p<0.0001), correlated significantly with cfDNA, especially in stage IV disease. Primary tumor site influenced cfDNA levels, with liver/bile duct cancers having higher cfDNA than other sites (median 13.5 vs 6.1 ng/mL, p<0.0001). Multivariable analysis confirmed overall tumor burden and hepatic tumor location (primary or metastatic) as principal drivers of cfDNA. Critically, cfDNA concentration was an independent predictor of shorter PFS (p=0.001) and DSS (p<0.0001), demonstrating increased value in advanced disease settings, irrespective of treatment intent.

**Conclusions:** CfDNA concentration is a robust, biologically integrated biomarker that provides an objective measure of total systemic tumor burden and prognosis in a large, pan-cancer cohort. By capturing disease activity irrespective of mutational status, it offers a valuable adjunct to targeted profiling, particularly in heterogeneous or advanced malignancies. Although these findings require prospective clinical validation, cfDNA concentration may eventually augment patient selection for aggressive versus palliative interventions, especially in advanced disease.

## Introduction

Liquid biopsy techniques have a growing range of clinical applications in surgical oncology, including prognostication, determining treatment eligibility, predicting response to neoadjuvant or adjuvant therapy, detecting minimal residual disease following curative-intent resection, and molecular surveillance for disease recurrence^1–4^. High-profile successes, such as the DYNAMIC study for circulating tumor DNA (ctDNA) guided adjuvant therapy in colon cancer, are expected to accelerate the uptake and integration of liquid biopsy techniques into management algorithms^5^.

However, the current landscape of commercial liquid biopsy assays is complex, with many methodologic variations, including sequencing platforms, gene panels, bioinformatics, and reporting metrics, which hinder comparisons across ctDNA assays^6–8^. Critically, these targeted approaches rely on the detection of specific clones, which may not represent the distinct biological behavior of the entire tumor bulk, particularly in the setting of spatial or temporal heterogeneity. Moreover, many assays require foreknowledge of patient-specific genomic alterations (tumor-informed assays) or the presence of a selected subset of common mutations. As such, these assays have limited utility for tumors with low mutational burden or rare mutations, tumors where biopsy is not feasible or safe, and tumors that are genomically heterogenous or subject to genetic drift during disease progression^9^. Assay reporting standards remain inconsistent and, in some cases, are reduced to a simplistic binary interpretation that may not capture the quantitative potential of liquid biopsy^10–12^. These operational and economic challenges remain major limiting factors for clinical uptake of liquid biopsy^13,14^.

Often overlooked is the raw plasma concentration of cell-free DNA (cfDNA) – the substrate for downstream sequencing applications. In contrast to ctDNA, total cfDNA concentration integrates shedding from all tumor sites, theoretically identifying biological overwhelm of clearance mechanisms, and capturing burden from clones that may be missed by targeted panels. We previously reported an institution-wide blood and solid tumor tissue biorepository in patients undergoing cancer care, focusing on the use of standardized protocols in blood collection and processing, along with a comprehensive clinicopathologic database^15^. In a pan-cancer series of 874 patients, we found enormous variability in the plasma concentration of input cfDNA, yet still found high concordance between plasma and solid tumor variants, even with small cfDNA amounts, indicating its accurate reflection of patient-specific cancer genomics^15^.

In this study we explored the utility of raw cfDNA concentration as a ‘molecular tumor burden’ biomarker by systematically quantifying its association with conventional clinical predictors and oncologic outcome in a pan-cancer cohort. Leveraging a unique repository of uniformly processed biospecimens and matched clinical data, our analysis investigated how cfDNA levels correlate with patient and tumor characteristics, particular anatomic sites of disease, and comprehensive measures of anatomic tumor burden, including tumor number, size, and metastatic patterns.

## Methods

### Consent and Sample Collection

This study enrolled adult cancer patients receiving clinical care at Allegheny Health Network (AHN) Cancer Institute in the setting of routine, clinical laboratory draws or prior to intravenous therapy. Detailed methods have been previously published and are summarized here^15^. All participants provided informed consent under an Allegheny Health Network Research Institute Institutional Review Board (IRB)-approved protocol (2020-258: Oncology Sample Biobank and Data Repository) allowing for the collection and de-identified use of their clinicopathologic and sequencing data. Whole blood was collected and maintained per manufacturer recommendations^15^. Samples were transported daily to the AHN Genomics Facility for processing, excluding those subjected to undue agitation, consistent with established guidelines^16,17^.

### Plasma Isolation and cfDNA Purification

Plasma was isolated via a previously described three-step sequential centrifugation protocol, designed for cell-free, debris-free plasma^15^. CfDNA was purified from thawed plasma, quantified with fluorometry, and fragments analyzed to delineate cfDNA distribution (75-300bp, 75-1200bp) and exclude germline DNA (1300-150000bp)^15,18^. The concentrations of these specific cfDNA components were calculated by multiplying the fluorometry quantitation by the respective fragment analysis percentages.

### Demographic and Clinical Variable Collection

A database of patient information was prospectively maintained for patients enrolled in the biobanking protocol. Patient age, sex, and primary tumor site were confirmed via manual electronic medical record (EMR) review. American Joint Committee on Cancer (AJCC) stage and treatment intent (explicitly stated or inferred by physician review) were obtained from clinician notes. Largest tumor dimension and number of discrete tumors at the time of the blood draw were determined from diagnostic imaging and pathology reports (with physician image review when necessary). Patients with diffuse metastatic patterns (malignant ascites, malignant pleural effusion, ‘innumerable’ liver or lung metastases, carcinomatosis, matted or diffuse lymphadenopathy, or cerebrospinal fluid involvement) were assigned a tumor number >20. In patients with metastatic disease, the location(s) of known disease at the time of blood draw were recorded. Time-to-event variables were collected from the time of blood draw and included progression-free survival (PFS) and disease-specific survival (DSS). Date of death was gathered from the EMR or open access sources; death was attributed to cancer unless EMR review indicated an unrelated cause. Progression was defined as clinical, radiographic, serologic, or pathologic evidence of disease progression via physician EMR review.

### Statistical Analysis

Associations between cfDNA concentration and variables were carried out using nonparametric tests as appropriate. Linear regression was carried using natural log-transformed values of right-skewed statistics. Survival analyses were carried out using Cox regression following natural log-transformation of right-skewed variables. Comparisons between categorical groups were assessed with the logrank test. Statistical analyses were performed with Stata v17.0 (StataCorp, LLC, College Station, TX, USA), R v4.1.2 (R Core Team) with RStudio v.2022.12.0+353 (Rstudio Team) or GraphPad Prism v10.6.1 (GraphPad Software). Statistical significance was defined as a two-tailed p-value <0.05.

## Results

Peripheral blood cfDNA concentration was determined from 1000 unique patient samples between 3/2/2021 and 2/4/2025 (**Supplemental Table 1**) with a wide range of ages (18-99 years, median=65) and even sex distribution (female=53%, male=47%). The most commonly represented malignancies were ovary/fallopian tube (10.6%), soft tissue sarcoma (9.3%), appendiceal cancer (7.8%), cutaneous melanoma (6.1%), breast cancer (5.7%), non-small cell lung cancer (5.5%) and uterine cancer (5.0%), with all other sites comprising <5% of the cohort (**Supplemental Table 1**). AJCC stages were proportionately represented (stage I, 18.9%, stage II, 12.3%, stage III, 27.5% and stage IV, 41.3%). At the time of blood collection, treatment intent was curative in 52%, palliative in 41%, adjuvant in 7%, and surveillance in 1%. During the follow-up period, 58% of patients achieved no evidence of disease (NED) status, and in a total survival analysis time of 1566.6 person-years, 290 deaths attributed to cancer and 451 progression events were observed.

CfDNA concentration varied across a broad dynamic range (median=6.25ng/mL; range=0.5-1132.9; IQR=3.9, 12.05) and was right-skewed (p<0.0001). Significant associations between cfDNA concentration and baseline patient and disease characteristics were seen, including patient age (rho=0.18, p<0.0001)(**Figure 1a**) and female sex (median 6.8 vs 5.9ng/mL, p=0.03)(**Figure 1b**). The latter effect was essentially unchanged in a sensitivity analysis excluding patients with breast, fallopian tube/ovary, uterine or prostate cancer, ruling out confounding effects of sex-specific cancers on this association (median 6.8 vs. 6.0ng/mL, p=0.09).

**Figure 1:**
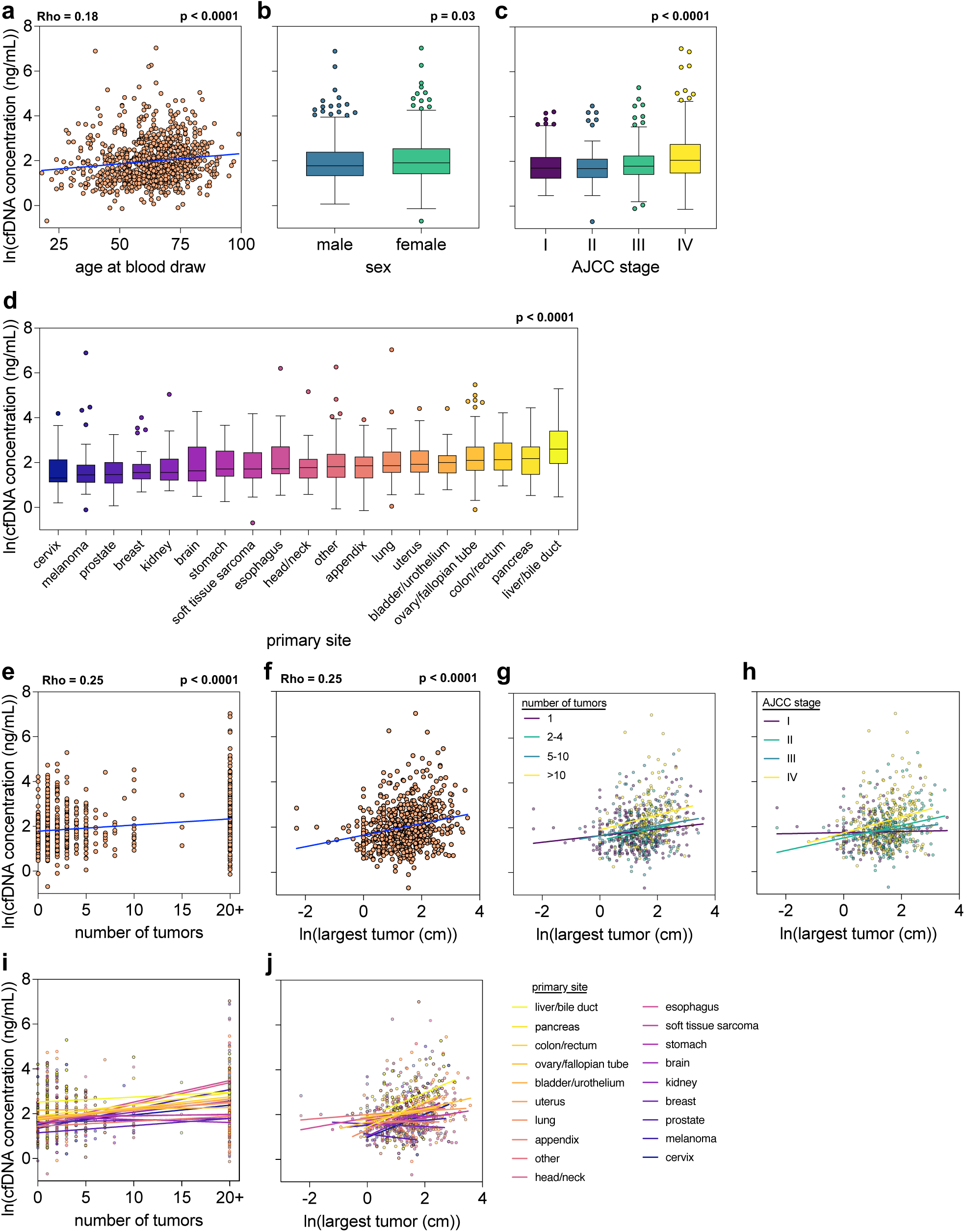
cfDNA concentration versus demographics, primary disease site, AJCC stage, and measures of anatomic tumor burden. a, Age versus cfDNA concentration as scatter plot with overlaid simple linear regression to illustrate linearity. b, Sex versus cfDNA concentration as box and whisker plots (Tukey’s method). c, AJCC stage versus cfDNA concentration as box and whisker plots (Tukey’s method). d, CfDNA concentration by primary site as box and whisker plots (Tukey’s method) in ascending order of median. e, Number of tumors versus cfDNA concentration as scatter plot with overlaid simple linear regression to illustrate linearity. f, Largest tumor diameters (cm, log transformed) versus cfDNA concentration as scatter plot with overlaid simple linear regression to illustrate linearity. g, Largest tumor diameters (cm, log transformed) versus cfDNA concentration as scatter plot with overlaid simple linear regression to illustrate linearity and with color stratification by number of tumors. h, Largest tumor diameters (cm, log transformed) versus cfDNA concentration as scatter plot with overlaid simple linear regression to illustrate linearity and with color stratification by AJCC stage. i, Number of tumors versus cfDNA concentration as scatter plot with overlaid simple linear regression to illustrate linearity and with color stratification by primary site. j, Largest tumor diameters (cm, log transformed) versus cfDNA concentration as scatter plot with overlaid simple linear regression to illustrate linearity and with color stratification by primary site. Statistical comparisons were performed using Spearman’s rank correlation with overlaid simple linear regression to illustrate linearity, Wilcoxon rank-sum test, Kruskal-Wallis test by ranks, or Jonckheere-Terpstra test for trend as appropriate. Natural log transformation of variables was performed as appropriate. CfDNA concentration given in ng/mL and natural log-transformed for visualization. AJCC: American Joint Committee on Cancer; cfDNA: cell-free DNA.

As previously reported, disease stage showed a tight association with cfDNA concentration (p<0.0001)(**Figure 1c**). CfDNA concentration also varied significantly across primary tumor sites (p=0.0001)(**Figure 1d)**. This variation persisted when the analysis focused only on the most common sites, mitigating potential influences from less common cancers with small sample sizes (p=0.0001). Notably, liver/bile duct cancers were associated with significantly higher cfDNA levels compared to every other primary tumor site, both as a pooled group and in individual site-specific comparisons (**Figure 1d**, **Table 1**). The median cfDNA concentration in liver/bile duct cancers was more than double that of other cancers (13.5 vs 6.1ng/mL, p<0.0001). This difference was consistent in both locoregionally-confined and metastatic cases.

**Table 1:**
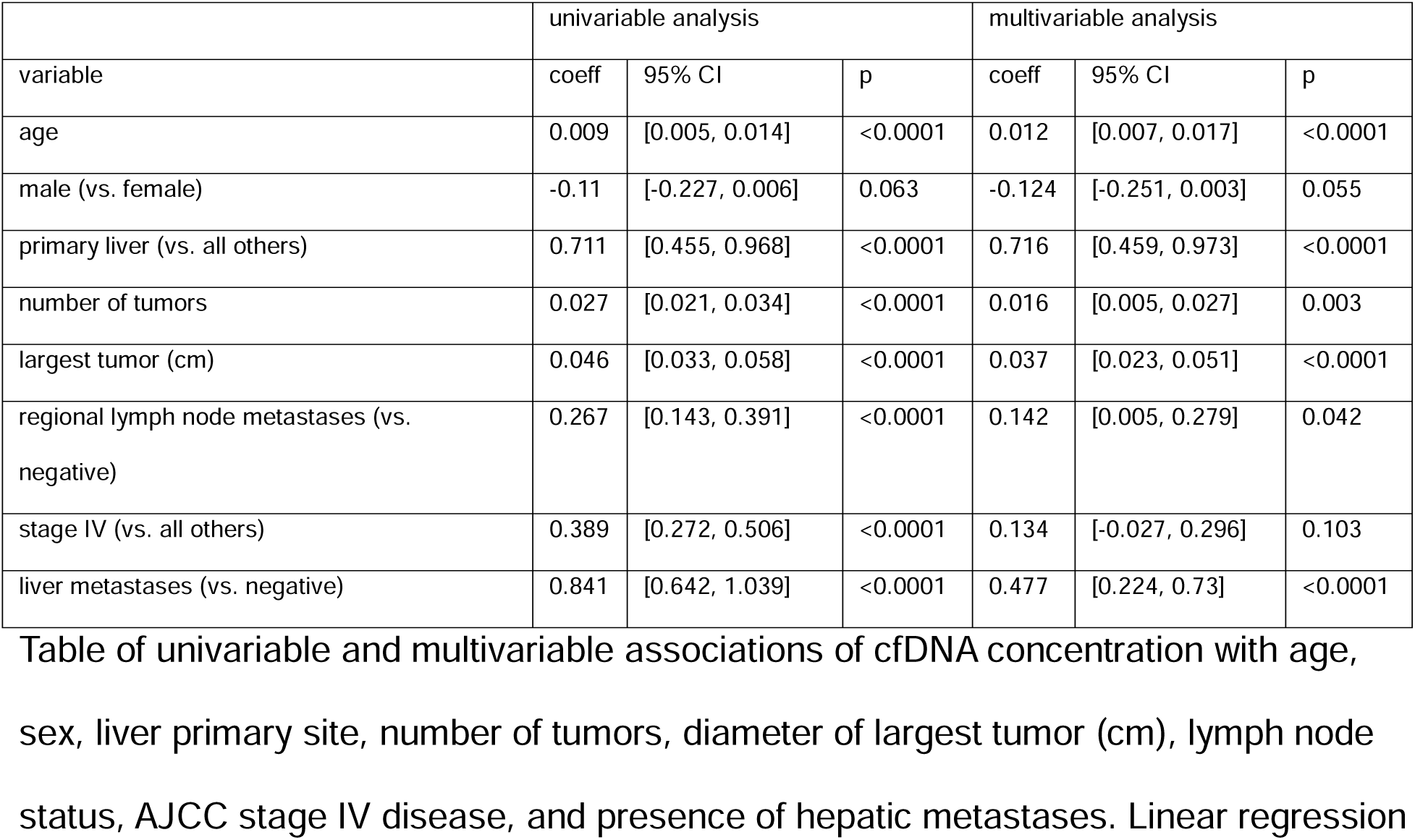
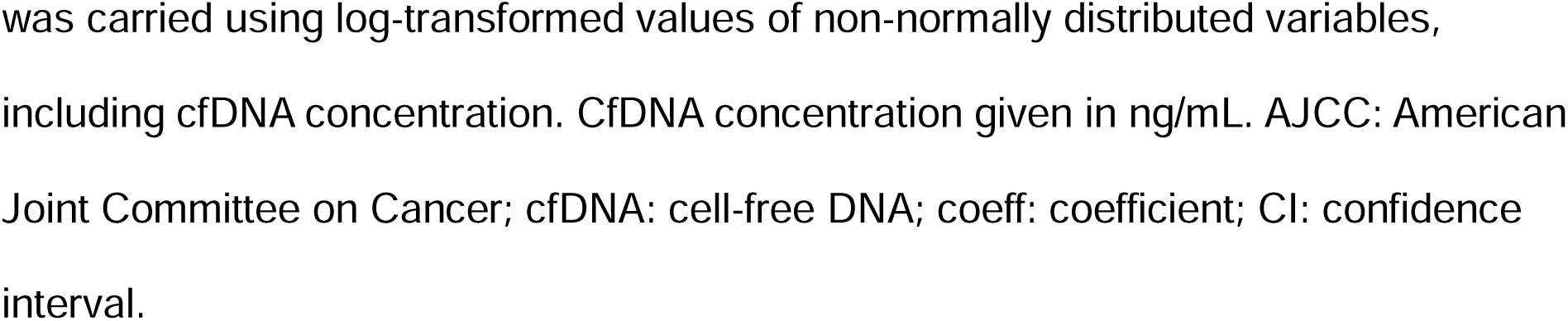
cfDNA concentration univariable and multivariable associations with patient characteristics.

| variable | univariable analysis |  |  | multivariable analysis |  |  |
| --- | --- | --- | --- | --- | --- | --- |
|  | coeff | 95% CI | p | coeff | 95% CI | p |
| age | 0.009 | [0.005, 0.014] | <0.0001 | 0.012 | [0.007, 0.017] | <0.0001 |
| male (vs. female) | -0.11 | [-0.227, 0.006] | 0.063 | -0.124 | [-0.251, 0.003] | 0.055 |
| primary liver (vs. all others) | 0.711 | [0.455, 0.968] | <0.0001 | 0.716 | [0.459, 0.973] | <0.0001 |
| number of tumors | 0.027 | [0.021, 0.034] | <0.0001 | 0.016 | [0.005, 0.027] | 0.003 |
| largest tumor (cm) | 0.046 | [0.033, 0.058] | <0.0001 | 0.037 | [0.023, 0.051] | <0.0001 |
| regional lymph node metastases (vs. negative) | 0.267 | [0.143, 0.391] | <0.0001 | 0.142 | [0.005, 0.279] | 0.042 |
| stage IV (vs. all others) | 0.389 | [0.272, 0.506] | <0.0001 | 0.134 | [-0.027, 0.296] | 0.103 |
| liver metastases (vs. negative) | 0.841 | [0.642, 1.039] | <0.0001 | 0.477 | [0.224, 0.73] | <0.0001 |

Table of univariable and multivariable associations of cfDNA concentration with age, sex, liver primary site, number of tumors, diameter of largest tumor (cm), lymph node status, AJCC stage IV disease, and presence of hepatic metastases. Linear regression
was carried using log-transformed values of non-normally distributed variables, including cfDNA concentration. CfDNA concentration given in ng/mL. AJCC: American Joint Committee on Cancer; cfDNA: cell-free DNA; coeff: coefficient; CI: confidence interval.

Assessing anatomic disease burden, we found a significant positive association between the number of discrete tumors and cfDNA concentration (p<0.0001) (**Figure 1e**). The cross-sectional diameter of the largest discrete tumor also showed a significant association with cfDNA across all cases (rho=0.25, p<0.0001). To minimize the influence of outliers, the analysis was repeated by limiting tumor size to <10cm (n=687, rho=0.18, p<0.0001), <5cm (n=500, rho=0.11, p=0.02), and <3cm (n=338, rho=0.12, p=0.03), confirming the stability of this effect across all tumor size ranges. This correlation was observed in both unifocal (n=290, rho=0.13, p=0.03) and multifocal disease (n=497, rho=0.31, p<0.0001) (**Figure 1f**).

Given the influence of tumor number, size, and disease stage on cfDNA, we investigated their interactions as surrogate markers of tumor burden. The association between largest tumor size and cfDNA concentration remained stable irrespective of tumor number, indicating its significance even with multiple tumors or widespread metastatic disease (**Figure 1g**). Surprisingly, the effect of largest tumor size on cfDNA was significant only in stage III (rho=0.28, p<0.0001) and stage IV (rho=0.27, p<0.0001) disease and not in localized disease (stage I, rho=0.03, p=0.68; stage II, rho=0.14, p=0.17), again highlighting a tight correlation between tumor size and cfDNA in the presence of multifocal or distant disease (**Figure 1h**), and challenging the prevailing emphasis on tumor size in the staging of localized disease.

To further characterize the influence of tumor size and number on cfDNA concentration, we next tested interactions between these factors and primary tumor site on cfDNA concentration. The size-cfDNA relationship varied across primary tumor sites, showing a moderate positive association in lung (rho=0.43, p=0.0002), liver/bile duct (rho=0.34, p=0.03), kidney (rho=0.28, p=0.09), pancreas (rho=0.28, p=0.07) and ovary/fallopian tube (rho=0.23, p=0.05) cancers, while other disease sites showed only weak (rho<0.2) associations (bladder/urothelium, cervix, uterus, soft tissue, melanoma, colon/rectum, esophagus, stomach, brain) or even negative associations (breast, rho=-0.05; head/neck, rho=-0.12; appendix, rho=-0.19), which did not meet statistical significance (**Figure 1i**). Likewise, the number of discrete tumors was strongly associated with cfDNA concentration in most disease sites, with moderate effect size for esophagus (rho=0.68, p<0.0001), bladder/urothelium (rho=0.53, p<0.006), prostate (rho=0.49, p=0.02), uterus (rho=0.47, p=0.0006), colon/rectum (rho=0.44, p=0.004), kidney (rho=0.31, p=0.05), head/neck (rho=0.26, p=0.1), melanoma (rho=0.24, p=0.07), liver/bile duct (rho=0.23, p=0.11), and appendix (rho=0.21, p=0.06) (**Figure 1j**). Collectively, these findings indicate that the site of primary tumor significantly influences the relationship anatomic disease burden and cfDNA concentration.

Given the substantial impact of primary tumor location on cfDNA concentration, we next investigated the influence of specific metastatic sites by comparing cfDNA levels in Stage IV patients with a particular metastatic site against those without that site. Liver (p<0.0001), pleural effusion (p=0.001), regional lymph nodes (p=0.0005), and non-regional lymph nodes (p=0.002) were associated with significantly increased cfDNA concentration relative to other metastatic sites (**Figure 2a**). The association of higher cfDNA in patients with regional lymph nodes in the presence of distant metastatic disease was surprising, given the prevailing assumption that distant disease ‘trumps’ regional disease. To further investigate this finding, we separately queried the effect of regional lymph node metastases in stage IV versus stage I-III patients. Interestingly, the effect of regional lymph node disease on cfDNA was observed only among stage IV patients (p<0.0001), and not among stage I-III patients (p=nonsignificant), implying that the nodal infiltration itself may not be the mechanism of cfDNA shedding into the blood (**Figure 2b**).

**Figure 2:**
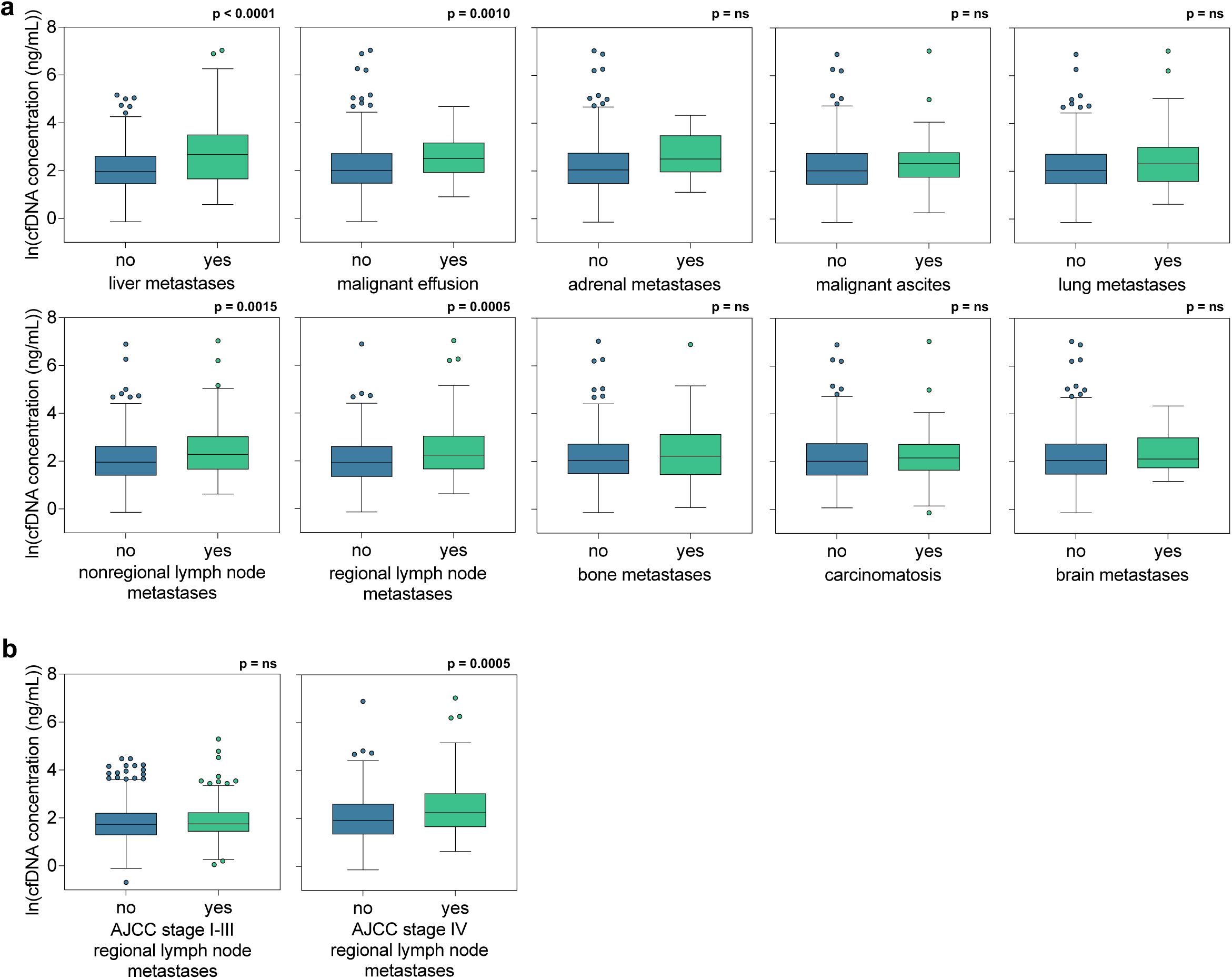
cfDNA concentration versus metastatic disease patterns, including sites of metastasis and regional lymph node status. a, Metastatic sites versus cfDNA concentration in stage IV patients as box and whisker plots (Tukey’s method). b, Regional lymph node metastasis status versus cfDNA concentration as box and whisker plots (Tukey’s method) stratified by AJCC stage I-III (left) and stage IV (right) disease. Statistical comparisons were performed using the Wilcoxon rank-sum test. CfDNA concentration given in ng/mL and natural log-transformed for visualization. AJCC: American Joint Committee on Cancer; cfDNA: cell-free DNA.

Next, we performed univariable and multivariable linear regression analyses incorporating predictors of cfDNA concentration to assess their relative importance and independence. In the univariable analysis, patient age, primary tumor site in the liver (vs. all others), number of tumors, diameter of the largest tumor, positive regional lymph nodes (vs. negative), Stage IV disease (vs. stages I-III), and presence of liver metastases (vs. absence) were all significantly associated with increased cfDNA concentration (p<0.05 for all). In the multivariable analysis, age, primary tumor site in the liver, number of tumors, diameter of the largest tumor, positive regional lymph nodes, and presence of liver metastases remained significant independent predictors of higher cfDNA concentration (p<0.05 for all). Notably, Stage IV disease was no longer statistically significant in the multivariable model, suggesting that its effect was largely accounted for by tumor burden (**Table 1**). These results suggest that overall tumor burden (size and number) and hepatic tumor location (primary and metastatic) were the principal drivers of cfDNA concentration in our cohort, although these effects may have been moderated by patient age and sex.

Univariable and multivariable Cox proportional hazards models were employed to identify factors associated with both progression-free survival (PFS) and disease-specific survival (DSS). In univariate analyses, higher cfDNA concentration, increased age, male sex (vs. female), larger diameter of the largest tumor, greater number of tumors, and advanced disease stage (Stage III and IV vs. Stage I) were all significantly associated with shorter PFS and DSS. Furthermore, most primary tumor sites, with breast cancer as the reference, also demonstrated significantly shorter survival in both univariate PFS and DSS analyses. Notably, in the multivariate models for both PFS and DSS, cfDNA concentration consistently emerged as a strong and highly significant independent predictor of shorter survival (PFS: p=0.001; DSS: p<0.0001)(**Table 2**). Kaplan-Meier survival curves constructed separately for Stage I-III and Stage IV patients visually convey the utility of cfDNA concentration for stratifying prognosis as measured by PFS and DSS, especially in patients with metastatic disease (**Figure 3a,b).**

**Figure 3:**
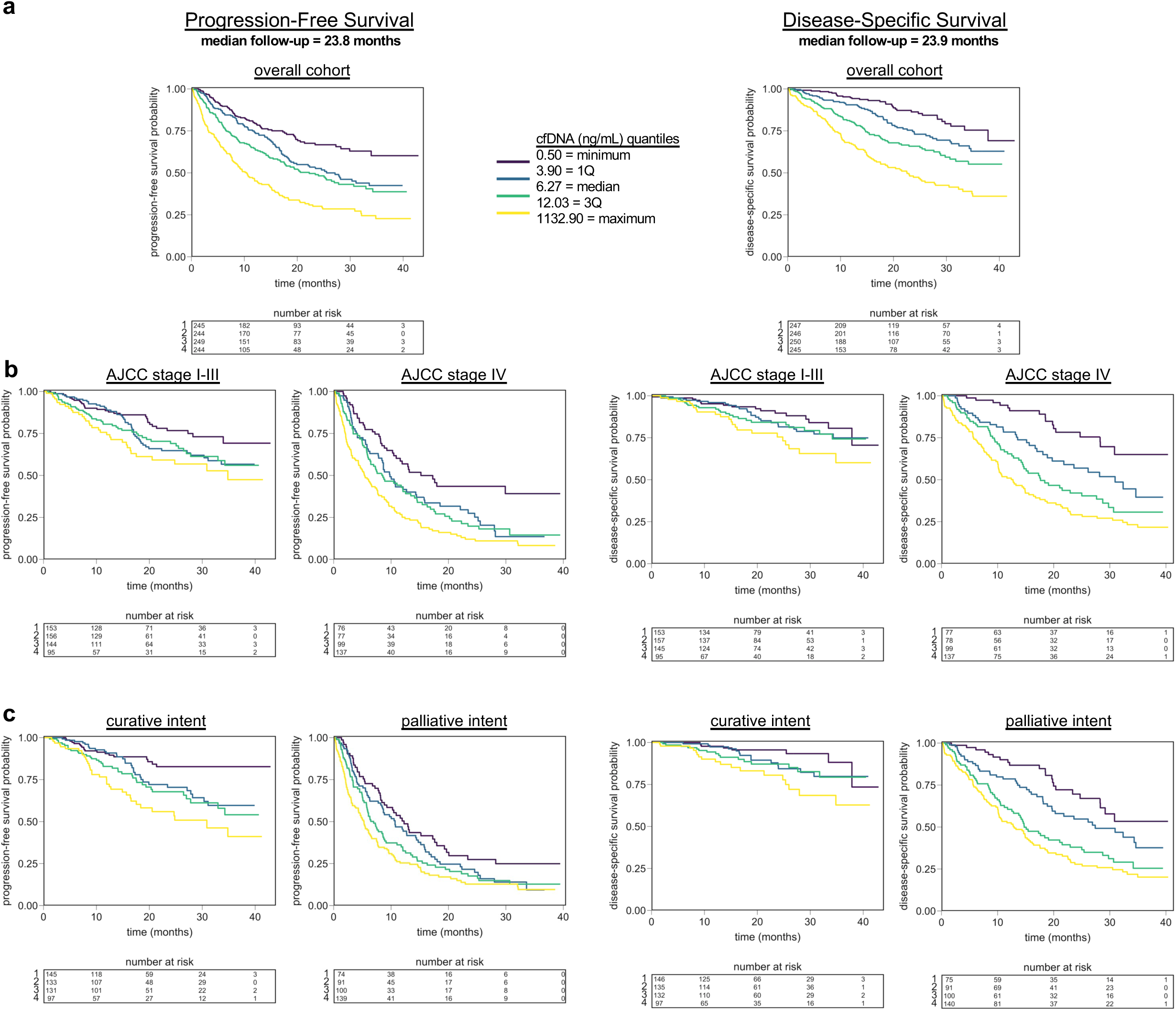
cfDNA concentration versus progression-free and disease-specific survival. a, PFS and DSS by cfDNA concentration quartiles (from low to high: 1, 2, 3, 4) in the overall cohort. b, PFS and DSS by cfDNA concentration quartile stratified by AJCC stage I-III (left) and stage IV (right) disease. c, PFS and DSS by cfDNA concentration quartile stratified by curative intent (left) and palliative intent (right). Time-to-event measured from time of blood draw. PFS is measured with disease progression as the event. DSS is measured with death from disease as the event. Comparisons between categorical groups were assessed with the logrank test. CfDNA concentration given in ng/mL. AJCC: American Joint Committee on Cancer; cfDNA: cell-free DNA; DSS: disease-specific survival; PFS: progression-free survival.

**Table 2:**
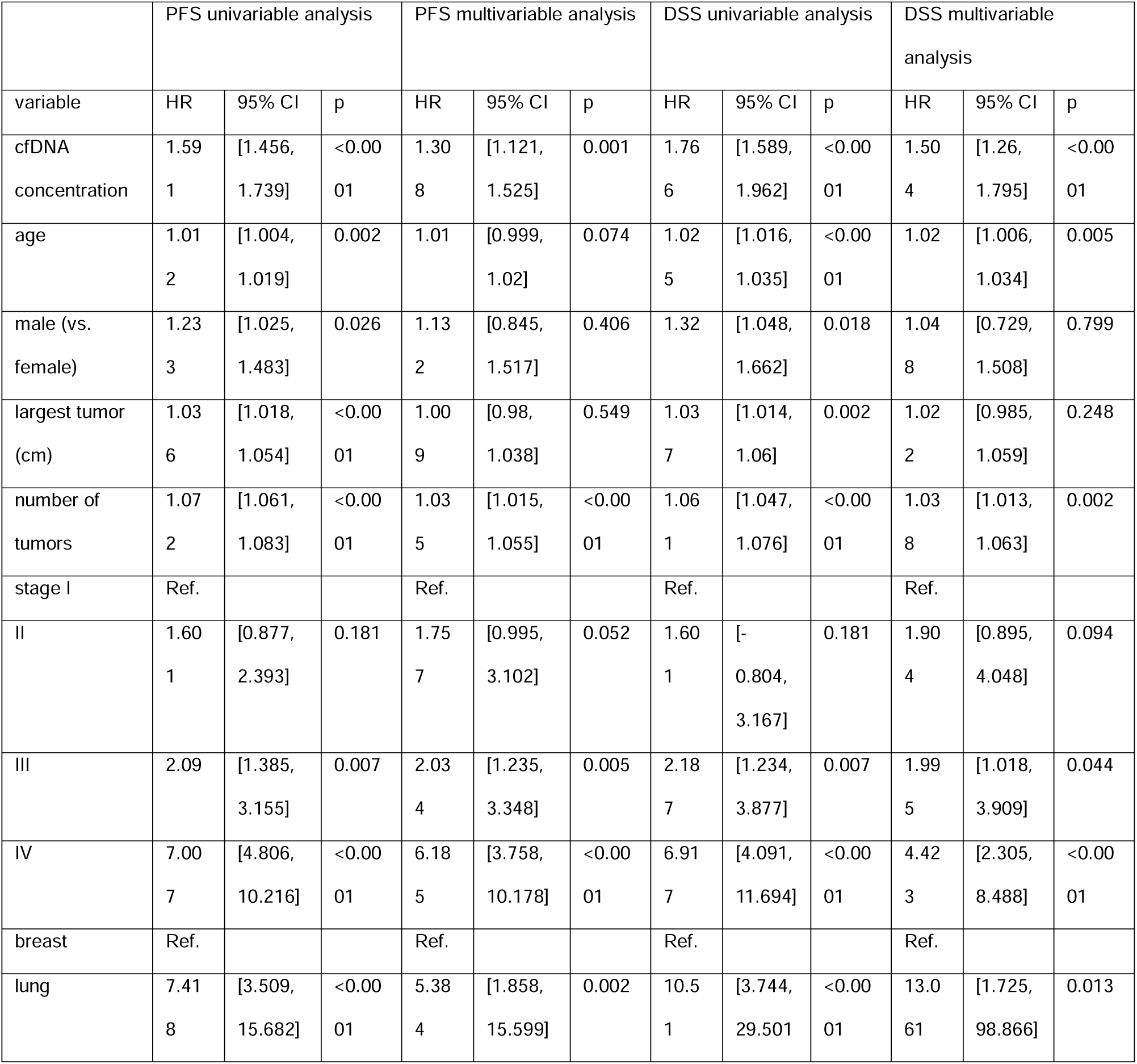
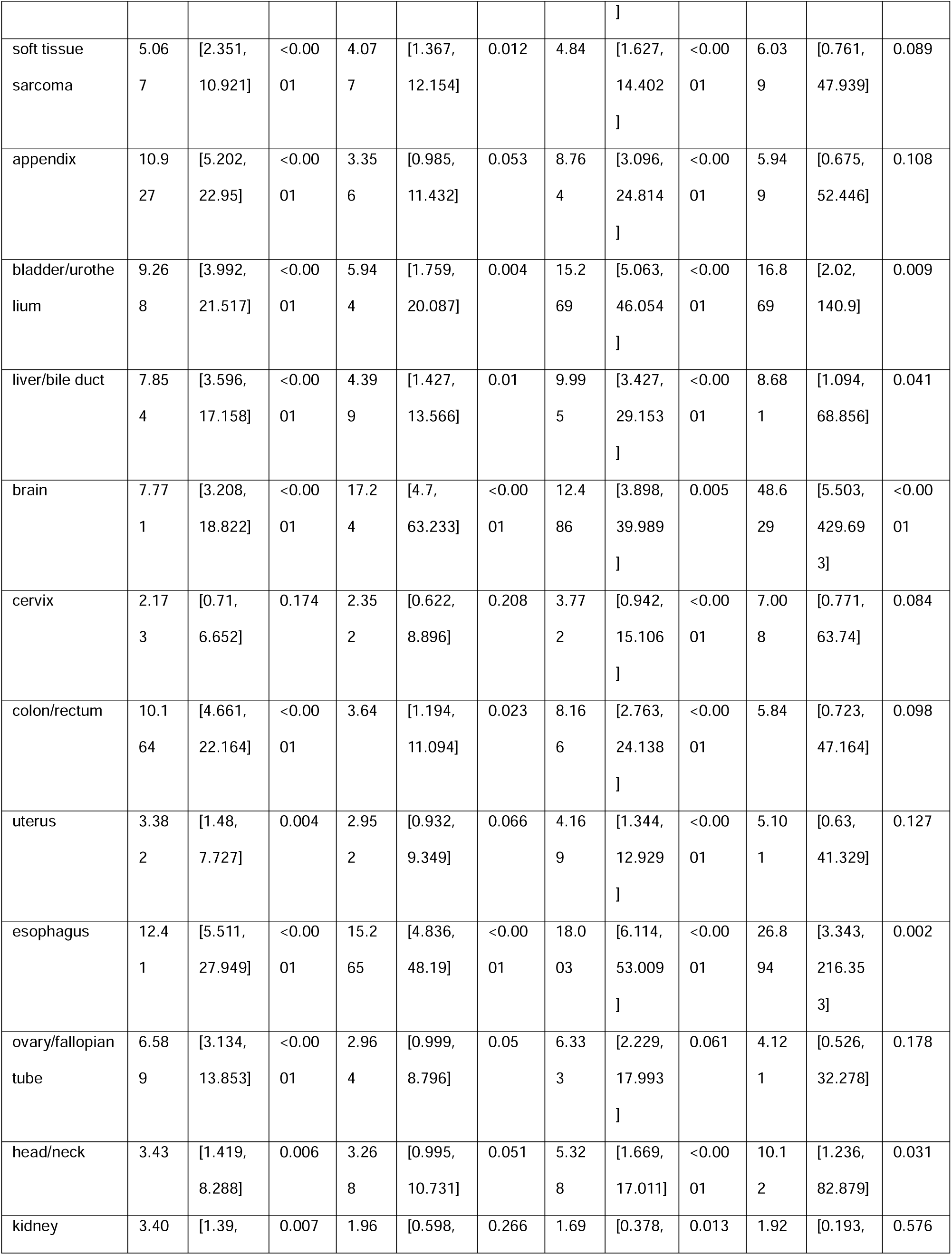
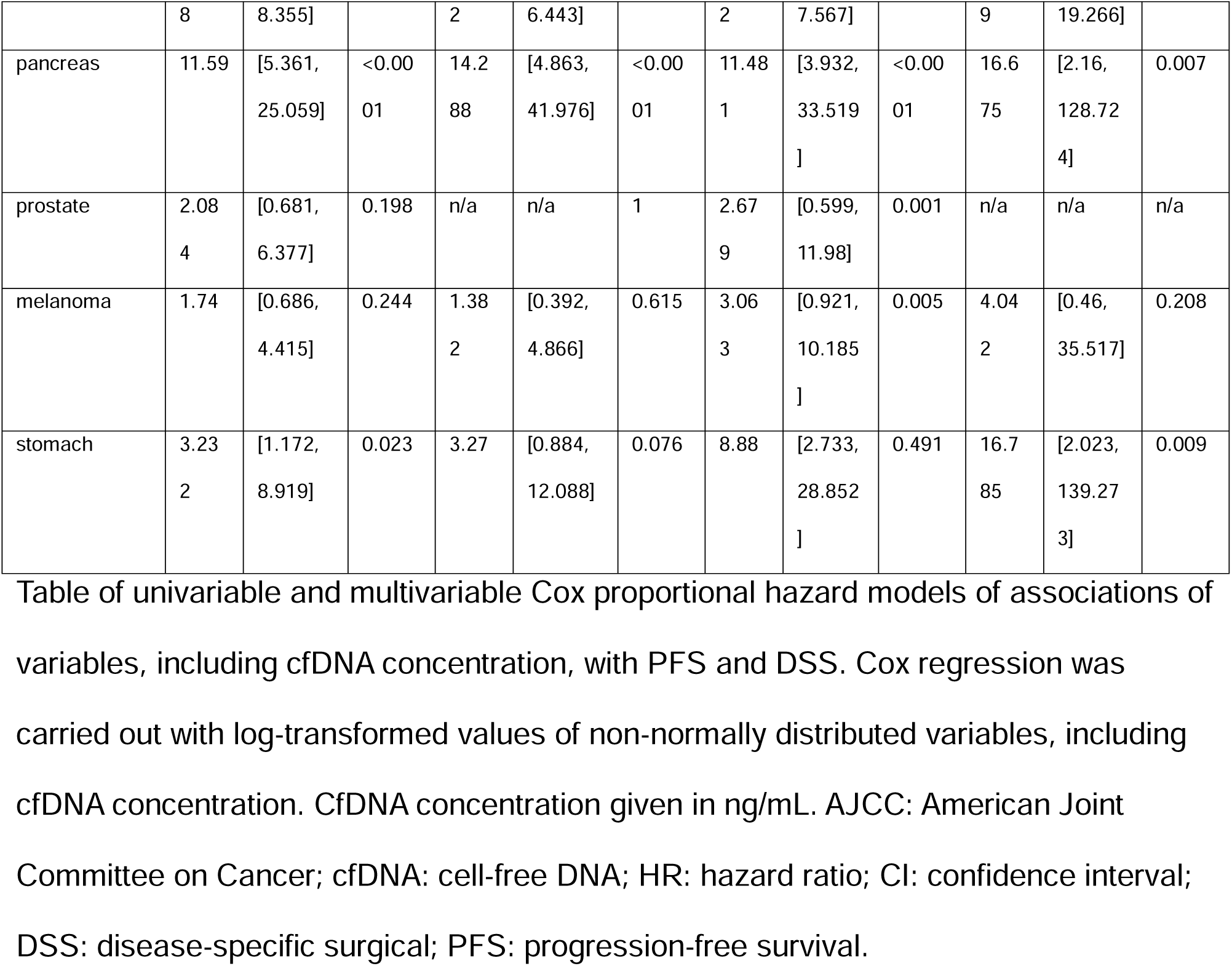
cfDNA concentration univariable and multivariable associations with PFS and DSS.

Finally, given the stable and independent influence of cfDNA concentration on prognosis, we used Cox regression for overall survival to assess whether these results were impacted by treatment intent (curative vs. surveillance vs. palliative). In this model, cfDNA concentration remained a highly significant independent predictor of shorter survival (p<0.0001). Regarding treatment intent, patients receiving palliative care had significantly shorter survival compared to those with curative intent (p<0.0001). Conversely, no statistically significant difference in survival was observed between patients under surveillance and those with curative intent (p=0.524). Importantly, in patients with stage IV disease undergoing curative-intent therapy (n=63), cfDNA concentration remained an extremely robust predictor of overall (HR=2.23, 95% CI=[1.05, 4.71], p=0.036) and progression-free survival (HR=2.65, 95% CI=[1.58, 4.27], p<0.0001). These findings reinforce the consistent prognostic utility of cfDNA concentration for survival, irrespective of treatment intent (**Figure 3c**; **Table 2**).

## Discussion

In this pan-cancer analysis, cell-free DNA concentration was significantly associated with a range of patient and tumor characteristics. The most pronounced determinants of cfDNA levels were related to anatomic tumor burden and liver involvement (either primary or secondary disease), suggesting a unique anatomic or physiological context influencing cfDNA release or clearance in these malignancies. Interestingly, the strong correlation between cfDNA levels and increasing tumor size was particularly evident in advanced stages with distant metastases and numerous sites of disease. Notably, cfDNA concentration emerged as a robust independent prognostic for both PFS and DSS, even after adjusting for AJCC stage and treatment intent. These results collectively suggest significant potential for this simple biomarker to assist in clinical management of solid tumor patients.

The role of circulating tumor DNA in managing solid tumors is expanding, as evidenced by several recent studies showing its utility in patient selection for adjuvant chemotherapy^5^, early response indication^19^, and treatment failure detection^20^. Notably, these and other ctDNA applications rely on DNA sequencing and mutation calling, both of which quantitation of cfDNA – the substrate for ctDNA – does not require. Circulating cfDNA originates from various sources, including hematopoietic lineages, vascular endothelial cells, neurons, hepatocytes, and, in cancer patients, tumor cells^21–23^. Normally an equilibrium of cfDNA level is maintained via rapid degradation by circulating enzymes like DNase I and clearance by the liver, spleen, and kidneys^24–26^. In cancer patients, increased cell death, active tumor secretion, and overwhelmed clearance mechanisms result in cfDNA accumulation^26–29^.

Conventionally, tumor burden has been estimated through a combined assessment of symptoms, physical exam or endoscopic findings, cross-sectional imaging, and serologic markers. Each of these modalities carry limitations, such as subjectivity, invasiveness, cost, radiation exposure, specificity/sensitivity balance, and lag time between the onset of treatment and tangible evidence of response or non-response. Our study demonstrates the robust nature of cfDNA to quantitatively assess patient tumor burden in a wide variety of cancers, stages, and metastatic patterns. This indicates the potential of cfDNA to circumvent many of these limitations by providing an objective, repeatable assessment.

The relationship between ctDNA concentration and tumor burden in prior studies, while generally correlative, exhibits considerable heterogeneity across cancer types, influenced by tumor biology, metastatic site, and non-tumor cfDNA sources^30–35^. Our analysis similarly revealed a relationship between anatomic tumor site and cfDNA concentration. Importantly, these data reinforce the liver’s mechanistic influence, previously suggested in animal models, within a pan-cancer cohort^26^. Our findings of a distinct elevation of cfDNA in hepatic disease suggests that this biomarker captures the physiological state of the clearance organs. Where mutation-specific assays might only detect the presence of a liver metastasis, total cfDNA concentration reveals the functional impact of that metastasis on systemic homeostasis, whether it be bypass or overwhelm of hepatic clearance. These findings underscore the importance of considering tumor location and metastatic pattern when assessing cfDNA concentration as a molecular biomarker of disease burden^36–38^.

The implications of our findings in the management of patients with metastatic disease warrant special consideration. In practice, aggressive treatments, including surgical locoregional interventions, are offered to selected patients with stage IV disease depending on clinician judgement of the potential for benefit. Selection criteria for these interventions remains highly subjective, with significant variation among institutions. In our series, cfDNA concentration remained highly predictive of progression-free and disease-specific survival when applied to the subset of patients with metastatic disease. We propose that plasma cfDNA concentration could substantially augment selection for invasive locoregional procedures. By providing a systemic tumor burden assessment that is immune to sampling error or mutational heterogeneity, cfDNA concentration helps identify patients whose systemic disease biology is too aggressive for local control, regardless of their anatomic resectability.

Despite its promise, the clinical application of cfDNA concentration carries limitations warranting careful consideration. The selection of assay and metric must be carefully tailored to the specific biological context and clinical question. Fragmentomic and methylation-based approaches may need to be incorporating to improve biomarker performance as these approaches mature^39–41^. Clarification is required as to whether non-tumor cfDNA sources (e.g., neutrophil extracellular traps, clonal hematopoiesis) affect biomarker performance^30,42^. As such, at present the interpretation of cfDNA concentration should integrate tumor anatomic site and assay limitations along with conventional (clinical and imaging) data to inform treatment decisions^40,41,43^. These findings require external validation in other practice settings, given they were obtained with stringent methods within a single institution, and prospective validation in the iterative disease-monitoring setting.

In summary, this study highlights cfDNA concentration as a promising, readily accessible biomarker that objectively reflects tumor burden and prognosis across a diverse range of cancers. Our findings underscore the critical influence of specific disease sites, particularly hepatic involvement, on cfDNA levels, suggesting its utility in refining patient assessment beyond traditional imaging. Ultimately, incorporating cfDNA concentration into clinical decision-making holds the potential to enhance patient stratification, guide treatment adjustments, and individualize management strategies more effectively.

## Abbreviations

ctDNA: circulating tumor DNA
cfDNA: cell-free DNA
IRB: institutional review board
AHN: Allegheny Health Network
EMR: electronic medical record
AJCC: American Joint Committee on Cancer
PFS: progression-free survival
DSS: disease-specific survival
NED: no evidence of disease

## Declarations

### Ethics Approval and Consent to Participate

All participants provided informed consent under an Allegheny Health Network Research Institute Institutional Review Board-approved protocol (2020-258: Oncology Sample Biobank and Data Repository) allowing for the collection and de-identified use of their clinicopathologic and sequencing data.

### Consent for Publication

Not applicable.

### Data Availability

The datasets used and/or analyzed during the current study are available from the corresponding author on reasonable request.

## Author Contributions

Concept and design: SLM, PLW

Acquisition, analysis, or interpretation of data: SLM, MC, CS, SP, NV, PP, PHG, PES, AHS, CJA, JMN, DLB, PLW

Drafting of the manuscript: SLM, PLW

Critical review of the manuscript for important intellectual content: SLM, AHS, CJA, JMN, TWR, OC, RSS, AHZ, WAL, DLB, PLW

Statistical analysis: SLM, PLW

Obtained funding: WAL, DLB, PLW

Administrative, technical, or material support: PP, PHG, PES, AHZ, WAL, DLB, PLW

Supervision: AHZ, WAL, DLB, PLW

## Acknowledgements

The authors thank the patients who participated in this study along with their families. They also wish to thank Tong Lam and Robin Barr who performed sample processing throughout the course of this study.

This work was selected as the first-place winner in the Basic Science Research category of the 2025 American College of Surgeons Commission on Cancer Paper

Competition and presented at the American College of Surgeons Commission on Cancer Plenary Session on October 4^th^, 2025.

