## Supplemental Table 1 for "Cell-Free DNA Concentration as a Mutation-Agnostic Readout of Systemic Tumor Burden and Prognosis: A Prospective Analysis of 1,000 Patients"

**Supplemental Tables and Legends**

**Supplemental Table 1: Summary of patient variables and associated cfDNA concentrations.**

| **continuous variables** | **rho** | **variable mean** | **variable median** | **variable minimum** | **variable maximum** |
| --- | --- | --- | --- | --- | --- |
| age (years) | 0.18 | 63.36 | 65.00 | 18.00 | 99.00 |
| number of tumors | 0.25 | 7.29 | 2.00 | 0.00 | 20.00 |
| largest tumor (cm) | 0.25 | 4.89 | 3.70 | 0.00 | 36.00 |
| **categorical variables** | **sublevel** | **sublevel frequency** | **sublevel proportion** | **sublevel cfDNA mean** |  |
| sex |  |  |  |  |  |
|  | female | 528 | 0.53 | 15.85 |  |
|  | male | 467 | 0.47 | 13.90 |  |
| stage |  |  |  |  |  |
|  | I | 179 | 0.19 | 8.50 |  |
|  | II | 116 | 0.12 | 8.82 |  |
|  | III | 260 | 0.27 | 10.34 |  |
|  | IV | 391 | 0.41 | 22.58 |  |
| intention of treatment |  |  |  |  |  |
|  | surveillance | 7 | 0.01 | 4.84 |  |
|  | adjuvant | 65 | 0.07 | 8.41 |  |
|  | curative | 513 | 0.52 | 9.59 |  |
|  | palliative | 406 | 0.41 | 22.90 |  |
| NED reached |  |  |  |  |  |
|  | no | 412 | 0.42 | 22.81 |  |
|  | yes | 570 | 0.58 | 9.35 |  |
| primary site |  |  |  |  |  |
|  | ovary/fallopian tube | 104 | 0.10 | 18.28 |  |
|  | other | 96 | 0.10 | 16.94 |  |
|  | lung | 81 | 0.08 | 23.55 |  |
|  | appendix | 81 | 0.08 | 8.72 |  |
|  | soft tissue sarcoma | 80 | 0.08 | 9.80 |  |
|  | melanoma | 59 | 0.06 | 24.65 |  |
|  | breast | 58 | 0.06 | 7.46 |  |
|  | uterus | 54 | 0.05 | 10.80 |  |
|  | liver/bile duct | 52 | 0.05 | 24.81 |  |
|  | pancreas | 50 | 0.05 | 13.02 |  |
|  | colon/rectum | 44 | 0.04 | 12.71 |  |
|  | head/neck | 41 | 0.04 | 10.79 |  |
|  | kidney | 41 | 0.04 | 11.76 |  |
|  | esophagus | 30 | 0.03 | 27.10 |  |
|  | brain | 29 | 0.03 | 11.02 |  |
|  | bladder/urothelium | 28 | 0.03 | 10.88 |  |
|  | stomach | 25 | 0.03 | 9.54 |  |
|  | cervix | 24 | 0.02 | 10.12 |  |
|  | prostate | 23 | 0.02 | 6.56 |  |
| metastatic disease |  |  |  |  |  |
|  | no | 555 | 0.59 | 9.43 |  |
|  | yes | 392 | 0.41 | 22.58 |  |
| malignant effusion |  |  |  |  |  |
|  | no | 927 | 0.93 | 14.75 |  |
|  | yes | 66 | 0.07 | 17.70 |  |
| malignant ascites |  |  |  |  |  |
|  | no | 902 | 0.91 | 13.49 |  |
|  | yes | 91 | 0.09 | 29.38 |  |
| carcinomatosis |  |  |  |  |  |
|  | no | 818 | 0.82 | 13.69 |  |
|  | yes | 175 | 0.18 | 20.82 |  |
| liver metastases |  |  |  |  |  |
|  | no | 906 | 0.91 | 10.73 |  |
|  | yes | 87 | 0.09 | 58.80 |  |
| lung metastases |  |  |  |  |  |
|  | no | 893 | 0.90 | 13.09 |  |
|  | yes | 100 | 0.10 | 31.46 |  |
| regional lymph node metastases |  |  |  |  |  |
|  | no | 680 | 0.68 | 11.90 |  |
|  | yes | 313 | 0.32 | 21.56 |  |
| nonregional lymph node metastases |  |  |  |  |  |
|  | no | 830 | 0.84 | 12.42 |  |
|  | yes | 163 | 0.16 | 27.69 |  |
| brain metastases |  |  |  |  |  |
|  | no | 965 | 0.97 | 14.99 |  |
|  | yes | 28 | 0.03 | 13.44 |  |
| bone metastases |  |  |  |  |  |
|  | no | 929 | 0.94 | 13.32 |  |
|  | yes | 64 | 0.06 | 38.09 |  |
| adrenal metastases |  |  |  |  |  |
|  | no | 980 | 0.99 | 14.87 |  |
|  | yes | 13 | 0.01 | 19.74 |  |

Demographics, anatomic tumor burden variables, and metastatic disease pattern variables are listed with frequencies, percentages, and mean cfDNA concentrations in ng/mL.
